# Confidence in government and rumors amongst international migrant workers involved in dormitory outbreaks of COVID-19: A cross-sectional survey

**DOI:** 10.1101/2021.07.08.21260237

**Authors:** Edina YQ Tan, Dalia Albarazi, Young Ern Saw, P Buvanaswari, Kinjal Doshi, Jean CJ Liu

## Abstract

**Background:** In the coronavirus disease (COVID-19) pandemic, confidence in the government and access to accurate information have been critical to the control of outbreaks. Although outbreaks have emerged amongst communities of international migrant workers worldwide, little is known about how they perceive the government’s response or their exposure to rumors.

**Methods:** Between 22 June to 11 October 2020, we surveyed 1011 low-waged migrant workers involved in dormitory outbreaks within Singapore. Participants reported their confidence in the government; whether they had heard, shared, or believed widely-disseminated COVID-19 rumors; and their socio-demographics. Logistic regression models were fitted to identify factors associated with confidence and rumor exposure.

**Results:** 1 in 2 participants (54.2%, 95% CI: 51.1-57.3%) reported that they believed at least one COVID-19 rumor. This incidence was higher than that observed in the general population for the host country (Singapore). Nonetheless, most participants (90.0%, 95% CI: 87.6-91.5%) reported being confident that the government could control the spread of COVID-19. Age was significantly associated with belief in rumors, while educational level was associated with confidence in government.

**Conclusions:** Our findings suggest that language and cultural differences may limit the access that migrant workers have to official COVID-19 updates. Correspondingly, public health agencies should use targeted messaging strategies to promote health knowledge within migrant worker communities.

## 1. Introduction

The coronavirus disease 2019 (COVID-19) pandemic has underscored how public cooperation is critical for containment strategies. For example, to reduce COVID-19 infection, the Centers for Disease Control and Prevention urged the public to wear face masks; undergo physical distancing; reduce travel; pursue vaccination; and subject themselves for testing, isolation and contact tracing [1]. While these measures are effective, successful implementation requires compliance from members of the public [2, 3].

A key aspect of compliance is whether the public trusts the government and has confidence in its COVID-19 response [4]. Across Europe, for example, adherence to health advisories was dampened in regions reporting low trust in policy-makers [5], or following national incidents where confidence was damaged [6]. When confidence is low, rumors also tend to be shared and believed in place of official communication [7]. For example, in April 2020, a rumor was spread in Iran that consuming alcohol can prevent COVID-19 infection [8]. In that month alone, 728 Iranians died from methanol poisoning – an eleven-fold increase from the year before [8]. In sum, these accounts underscore the centrality of public trust for managing COVID-19, and the need to understand how the community views the situation and their reliance on rumors [9].

Although several national surveys have described public confidence and belief in COVID-19 rumors [4, 10-13], these have largely involved convenient samples where special populations are under-represented. Of note, we know little about the views of international migrant workers – a population of 164 million individuals employed outside their countries of birth [14]. This is a group whose views need to be understood for several reasons. First, a large number of migrant workers are employed in low-waged, manual labour jobs involving high-density work (e.g., factories) or living arrangements (e.g., dormitories that host up to 25,000 residents). As physical distancing is difficult to achieve, several large-scale outbreaks have occurred amongst migrant worker groups in Singapore, Thailand, Malaysia, and the Gulf states [15-17]. Second, migrant workers often face barriers accessing healthcare, information, or resources in their host countries [18-20]. These may prevent them from receiving official COVID-19 updates, skewing their risk assessments or increasing their reliance on rumors [20].

To address the gap in the literature, we recorded the views of migrant workers in Singapore during the COVID-19 pandemic. The city state is widely considered a ‘high performing health system’ on account of its low case fatality rate (0.05%) and minimal movement restrictions [21]. Surveys of the general population allude to high public confidence in the government, and low rates of sharing or belief in COVID-19 rumors [4, 22, 23]. Despite these statistics, Singapore provides a case study of health inequalities. Of the 62,000 COVID-19 cases reported to date, 9 in 10 have arisen from the 400,000 male migrant workers employed in Singapore’s construction, shipping, and process sectors [17, 24]. One study reported a disease prevalence rate 188 times higher amongst workers living in dormitories (47%) than in the general community (0.25%) [25], and all workers have been placed under prolonged movement restrictions to contain the spread (April-August 2020: complete dormitory lockdown; August 2020-the present time: gradual resumption of work and limited recreation activities). Against this backdrop, we documented confidence in government and the spread of rumors amongst 1011 migrant workers.

## 2. Methods

### 2.1 Study population

Data were taken from the COVID-19 Migrant Health Study, a cross-sectional survey conducted in Singapore between 22 June to 11 October 2020 [26]. Respondents were male migrants employed in manual labour jobs, and were included if they met the following eligibility criteria: aged 21 and above, and holding a government-issued permit indicating their employment status. Recruitment took place in-person at: (1) a dormitory complex associated with the largest COVID-19 cluster, (2) transient accommodation for workers relocated from dormitories, (3) a construction work site, and (4) a recreation centre for migrant workers. Additionally, an online survey link was advertised through physical posters and on messaging groups (WhatsApp and Telegram) at: (1) quarantine sites for active COVID-19 cases, and (2) worker dormitories. Surveys were made available in the primary languages spoken by migrant workers (English, Bengali, Tamil, Mandarin), and included both audio recordings and written text to ensure survey access regardless of literacy level.

All participants provided informed consent in accordance with a protocol approved by the Institutional Review Boards of the National University of Singapore and Singapore Health Services (ClinicalTrials.gov registration: NCT04718519). Participants who were recruited in-person received SGD $10 for their time.

### 2.2 Survey development

Following analogous surveys conducted during the Ebola and COVID-19 outbreaks [22, 27], we measured public confidence with three questions: (1) how confident participants were that the government could control the nationwide spread of COVID-19 (4-point scale ranging from “Not confident at all” (1) to “Very confident” (4)); (2) how fearful they were about their health during the COVID-19 situation (4-point scale ranging from “Not scared at all” (1) to “Very scared” (4)); and (3) how fearful they were about losing their job during this period (4-point scale ranging from “Not scared at all” (1) to “Very scared” (4)). Participants who had not tested positive for COVID-19 were also asked to judge the likelihood that they would be infected (4-point scale ranging from “Not at all likely” (1) to “Very likely” (4)).

To assess rumor spread, participants also indicated whether they had heard, believed, or shared each of the following COVID-19 rumors (yes / no): (1) drinking water frequently will help prevent infection (COVID-19 prevention); (2) eating garlic can help prevent infection (COVID-19 prevention); (3) the outbreak arose from people eating bat soup (COVID-19 origins); (4) the virus was created in a US lab to affect China’s economy (COVID-19 origins); and (5) the virus was created in a Chinese lab as a bioweapon (COVID-19 origins). On a global scale, these rumors have been widely disseminated and have been studied in other surveys [23]. To provide further context, we also asked participants to estimate how much time they spent each day: (i) looking for updates about COVID-19 (e.g., searching and reading news, browsing websites, watching videos), and (ii) using social media (e.g., WhatsApp, Facebook, TikTok) to discuss or share information about COVID-19.

Finally, we obtained the following sociodemographic data: age, country of origin (Bangladesh, India, Others), marital status (married, not married: single/widowed/separated/divorced), education (primary, secondary, tertiary), years spent in Singapore (≤ 5 years, > 5 years), and history of COVID-19 (tested positive: yes, no).

### 2.3 Statistical analysis

We first summarised public confidence and the spread of rumors (hearing, believing, and sharing rumors) using counts (%) and means. Where comparisons were made with other national surveys, we ran tests for equality of proportions with continuity corrections (for public confidence) or Welch’s t-tests (for rumor spread).

Binary logistic models were run to identify socio-demographic predictors of participants’: confidence in the government (confident: ‘very confident’ and ‘somewhat confident’; not confident: ‘not very confident’ and ‘not confident at all’) [Model 1]; exposure to rumors (low exposure: 0-2 rumors; high exposure: 3-5 rumors) [Model 2]; likelihood of believing rumours (believed at least one rumour, did not believe any rumours) [Model 3]; and likelihood of sharing rumours (shared at least one rumour, did not share any rumours) [Model 4]. Each model involved the following set of predictors: age, country of origin (Bangladesh, India, others), marital status (married, not married: single/widowed/separated/divorced), education (primary, secondary, tertiary), years spent in Singapore (≤ 5 years, > 5 years), and history of COVID-19 (tested positive: yes, no).

Bonferroni correction was applied to each model to control the Type 1 family-wise error rate at 0.05 (Bonferroni-adjusted alpha level of 0.05/8 predictors = 0.006). Analyses were conducted using SPSS Version 20 and R Version 4.0.3.

## 3. Results

### Baseline participant characteristics

We included data from 1011 survey respondents (78.5% response rate: 87.2%, 882/1011 in-person recruitment; 12.8%, 129/1011 online recruitment). 19.6% (198/1011) were sampled during complete quarantine restrictions (confined to the dormitory), 15.5% (157/1011) under moderate restrictions (confined to the dormitory and work sites), and 64.9% (656/1011) under minimal restrictions (confined to the dormitory, work sites, and recreation centres for leisure).

As shown in Table 1, participants were men with a mean age of 33.2 years (SD: 6.7) and came primarily from South Asia (Bangladesh: 57.1%, 577/1010; India: 37.8%, 382/1010). The majority had spent >5 years in Singapore (63.8%, 643/1008), were married (62.7%, 634/1011), and had at least secondary levels of education (89.4%, 896/1002). 1 in 3 participants had been diagnosed with COVID-19 (35.7%, 360/1009).

**Table 1.** *Baseline characteristics of respondents* (N = 1011)

|  | **M (SD) or n (%)** |
| --- | --- |
| **Demographic** |  |
| Gender | 1011 (100) |
| Age | 33.2 (6.7) |
| Ethnicity |  |
| Bangladesh | 577 (57.1) |
| India | 382 (37.8) |
| Others | 51 (5.0) |
| Marital status |  |
| Not Married | 377 (37.3) |
| Married | 634 (62.7) |
| Education |  |
| Primary | 106 (10.6) |
| Secondary | 493 (49.2) |
| Tertiary | 403 (40.2) |
| Years in Singapore |  |
| ≤ 5 years | 365 (36.2) |
| > 5 years | 643 (63.8) |
| Tested positive for COVID-19 |  |
| No | 649 (64.3) |
| Yes | 360 (35.7) |
| **Public confidence** |  |
| Confidence in government |  |
| Not confident at all | 79 (7.8) |
| Not very confident | 22 (2.2) |
| Somewhat confident | 114 (11.3) |
| Very confident | 793 (78.7) |
| Fear for health |  |
| Very scared | 76 (7.5) |
| Somewhat scared | 323 (32.0) |
| Not very scared | 125 (12.4) |
| Not scared at all | 484 (48.0) |
| Fear for job |  |
| Very scared | 132 (13.1) |
| Somewhat scared | 274 (27.2) |
| Not very scared | 99 (9.8) |
| Not scared at all | 503 (49.9) |
| Likelihood of COVID-19 infection |  |
| Very likely | 54 (8.3) |
| Somewhat likely | 81 (12.5) |
| Not too likely | 110 (17.0) |
| Not at all likely | 403 (62.2) |
| **Rumor spread** |  |
| Time spent checking COVID-19 news (hrs/day) | 1.98 (2.83) |
| Time spent discussing COVID-19 on social media (hrs/day) | 1.86 (2.48) |
| Rumor exposure |  |
| Low exposure (0-2 rumors) | 474 (46.9) |
| High exposure (3-5 rumors) | 537 (53.1) |
| Believed rumors |  |
| Did not believe any rumors | 463 (45.8) |
| Believed at least one rumor | 548 (54.2) |
| Shared rumors |  |
| Did not share any rumors | 845 (83.6) |
| Shared at least one rumor | 166 (16.4) |

### 3.2 Public confidence

9 in 10 participants reported confidence that the government could control the spread of COVID-19 (90.0% very or somewhat confident; 95% CI: 87.6-91.5%). Correspondingly, the majority of participants reported low levels of fear for their health (60.4% not scared at all or not very scared; 95% CI: 57.3-63.4%) or of losing their job during the pandemic (59.7% not scared at all or not very scared; 95% CI: 56.6-62.8%). Amongst those who had not previously tested positive, most judged it unlikely that they would be infected with COVID-19 (79.2% not at all likely or not too likely; 95% CI: 75.8-82.2%).

Applying binary logistic modelling, we found that education level significantly predicted confidence in government (Table 2). Relative to participants with primary education, participants with secondary or tertiary educational levels were more likely to report confidence in the local government (secondary: adjusted odds ratio [AOR] = 2.26, 95% CI: 1.26-4.04; tertiary: AOR = 2.50, 95% CI: 1.33-4.73).

**Table 2.** Predicting confidence in government and rumor spread among migrant workers during the COVID-19 outbreak.

|  | **Model 1** |  | **Model 2** |  | **Model 3** |  | **Model 4** |  |
| --- | --- | --- | --- | --- | --- | --- | --- | --- |
|  | **(Confidence in government)** |  | **(Rumor exposure)** |  | **(Rumor belief)** |  | **(Rumor sharing)** |  |
| **Variable** | **AOR (95% CI)** | ***p*** | **AOR (95% CI)** | ***p*** | **AOR (95% CI)** | ***p*** | **AOR (95% CI)** | ***p*** |
| Age | 1.00 (0.96, 1.05) | 0.88 | 1.00 (0.97, 1.02) | 0.78 | 1.05 (1.02, 1.07) | 0.001* | 1.00 (0.97, 1.04) | 0.80 |
| Ethnicity |  |  |  |  |  |  |  |  |
| Bangladesh | 0 (ref) |  | 0 (ref) |  | 0 (ref) |  | 0 (ref) |  |
| India | 1.35 (0.79, 2.33) | 0.27 | 1.50 (1.01, 2.06) | 0.01 | 1.32 (1.00, 1.81) | 0.08 | 1.58 (1.04, 2.41) | 0.03 |
| Others | 0.77 (0.31, 1.89) | 0.57 | 1.65 (0.89, 3.06) | 0.11 | 0.97 (0.52, 1.79) | 0.91 | 2.62 (1.29, 5.34) | 0.008 |
| Marital status |  |  |  |  |  |  |  |  |
| Not Married | 0 (ref) |  | 0 (ref) |  | 0 (ref) |  | 0 (ref) |  |
| Married | 0.96 (0.58, 1.61) | 0.89 | 0.94 (0.69, 1.28) | 0.71 | 0.90 (0.66, 1.22) | 0.50 | 0.81 (0.53, 1.23) | 0.32 |
| Education |  |  |  |  |  |  |  |  |
| Primary | 0 (ref) |  | 0 (ref) |  | 0 (ref) |  | 0 (ref) |  |
| Secondary | 2.26 (1.26, 4.04) | 0.006* | 1.23 (0.80, 1.89) | 0.35 | 0.61 (0.39, 0.94) | 0.03 | 0.57 (0.33, 0.99) | 0.04 |
| Tertiary | 2.50 (1.33, 4.73) | 0.005* | 1.14 (0.73, 1.79) | 0.57 | 0.71 (0.45, 1.13) | 0.15 | 0.88 (0.51, 1.52) | 0.64 |
| Years in Singapore |  |  |  |  |  |  |  |  |
| ≤ 5 years | 0 (ref) |  | 0 (ref) |  | 0 (ref) |  | 0 (ref) |  |
| > 5 years | 1.09 (0.67, 1.78) | 0.74 | 0.74 (0.55, 1.00) | 0.05 | 0.76 (0.57, 1.02) | 0.07 | 0.63 (0.42, 0.93) | 0.02 |
| Tested positive for COVID-19 |  |  |  |  |  |  |  |  |
| No | 0 (ref) |  | 0 (ref) |  | 0 (ref) |  | 0 (ref) |  |
| Yes | 0.84 (0.53, 1.33) | 0.45 | 1.12 (0.84, 1.49) | 0.43 | 1.00 (0.75, 1.34) | 0.98 | 1.16 (0.78, 1.73) | 0.45 |

*Note:* AOR=adjusted odds ratio; CI=confidence interval; \*indicates significance at $p < 0.006$ (after Bonferroni correction)

### 3.3 Exposure to COVID-19 rumors

Participants spent an average of 1.98 hours (SD = 2.83) each day looking for COVID-19 news, and 1.86 hours (SD = 2.48) using social media to discuss or share COVID-19 content. Against this backdrop, 88.9% (95% CI: 86.6-90.8%) of participants had heard at least one COVID-19 rumor (mean exposure: 2.64 rumors, SD = 1.53), with the most widely-heard rumor being that drinking water frequently can prevent infection (rate of hearing: 70.5%, 95% CI: 67.6-73.3%) (Figure 1). No socio-demographic variable significantly predicted rumor exposure in logistic regression analyses (Table 2).

**Figure 1.**
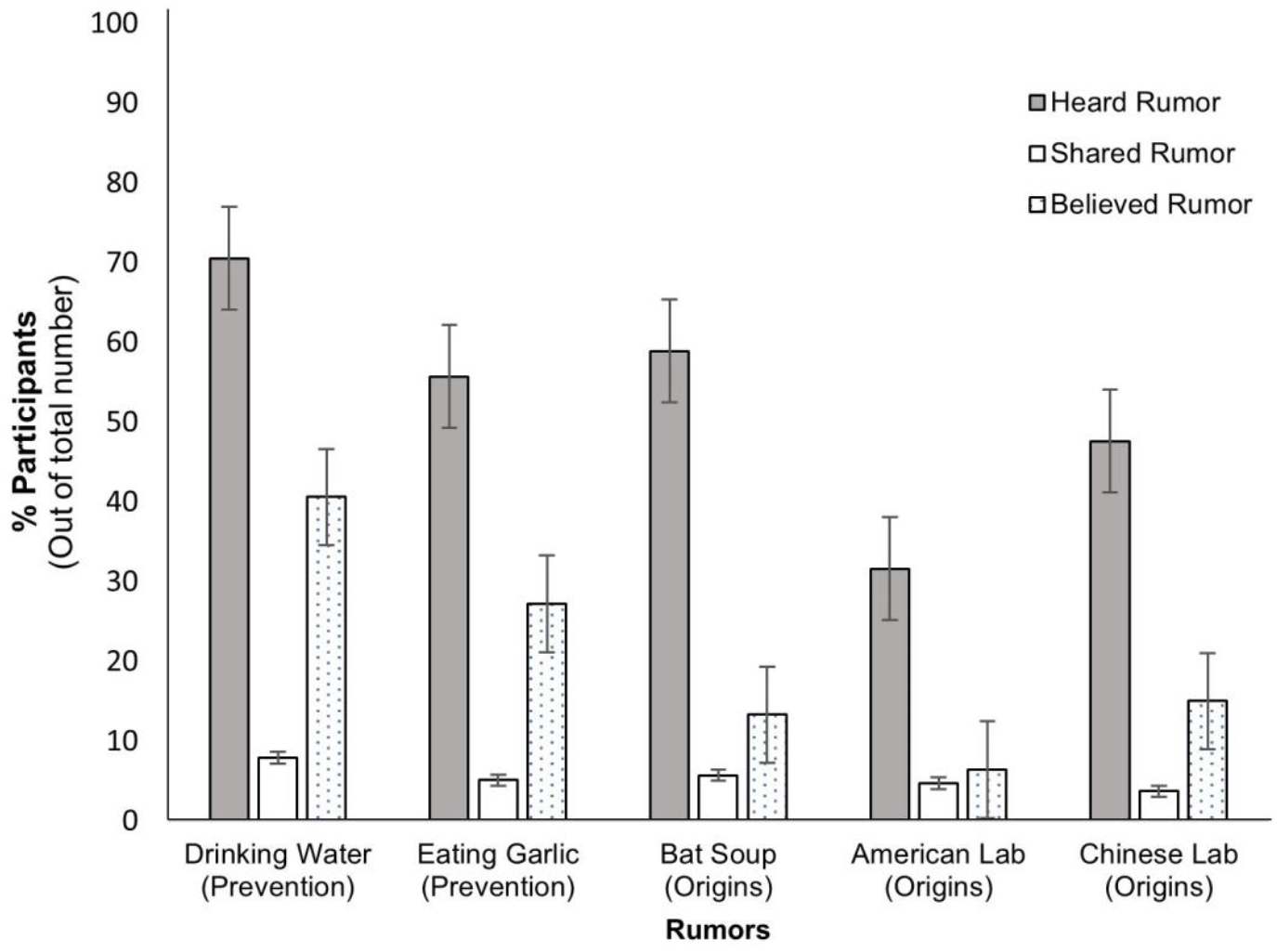
Percentage of participants hearing, sharing, or believing widely-disseminated COVID-19 rumors (that drinking water or eating garlic can prevent infection; or that the virus originated from bat soup consumption, an American lab, or a Chinese lab). Vertical lines represent 95% confidence intervals.

### 3.4. Believing and sharing COVID-19 rumors

Belief in COVID-19 rumors was high amongst migrant workers, with 1 in 2 participants (54.2%, 95% CI: 51.1-57.3%) reporting that they believed at least one rumor (mean rumors believed = 1.02, SD = 1.24). Again, the most widely believed rumor was that drinking water was a preventive measure (rate of belief: 40.6%, 95% CI: 37.5-43.7%) (Figure 1). When we applied logistic regression, older participants were more likely to report belief in rumors (AOR = 1.05, 95% CI: 1.02-1.07; Table 2).

Finally, few participants (16.4%, 95% CI: 14.2-18.9%) shared COVID-19 rumors with others (mean rumors shared = 0.27, SD = 0.74). Overall rates of rumor-sharing were similar across rumors (range: 3.7% to 7.9%), and no variable emerged as a significant predictor of sharing behavior in logistic regression models (Figure 1 and Table 2).

### 3.5 Comparisons with the local resident population

As a context, we compared participants’ responses to earlier community surveys that had been conducted within the Singapore resident population [22, 23]. Relative to the wider population, a higher percentage of migrant workers: (i) were confident that the government could control the spread of COVID-19 (90.0% vs. 86.2%; χ^2^[1, *N* = 2071] = 6.76, *p* = 0.009), and (ii) judged it unlikely that they would be infected (79.2% vs. 60.9%; χ^2^[1, *N* = 1711] = 60.93, *p* < 0.001). Although migrant workers had heard significantly fewer rumors (mean exposure: 2.64 vs. 3.34; *t*(2014.8) = 11.44, *p* < 0.001), they believed and shared more rumours (mean rumors believed: 1.02 vs 0.27, *t*(1379.7) = -17.67, *p* < 0.001; mean rumors shared: 0.27 vs. 0.18, *t*(1990.3) = -3.06, *p* = 0.002).

## 4. Discussion

In this study, we described for the first time confidence in government and the spread of rumors amongst migrant workers involved in COVID-19 outbreaks. As similar clusters have arisen amongst migrant communities worldwide, this line of work is critical from both a public health and humanitarian standpoint.

As our first observation, we found that participants believed and shared more COVID-19 rumors than the general population. This pattern of data may have arisen for several reasons. First, commentators have voiced concerns about migration-related barriers (e.g., language, literacy, social barriers) that may limit access to official COVID-19 updates [20]. If access is limited, rumors may then be turned to as a substitute source for information. Alternatively, rumors also tend to be spread during anxiety-provoking situations [28]. Given that migrant workers have faced the brunt of infection clusters and quarantine protocols [25], it follows that rumors could be spread more readily in these settings.

Beyond the quantity of rumors circulated, we also observed that the *nature* of rumors heard, shared, and believed differed between migrant workers and the resident population. For example, the most widely-heard rumor in our sample was that drinking water prevents infection. This same rumor was the least heard in the resident population, who instead reported high exposure to conspiratorial rumors (e.g., that COVID-19 originated from bat soup, an American lab, or a Chinese lab [23]). Given differing patterns of rumor exposure, our findings make a strong case that risk communication should be tailored for migrant worker communities. Notably, additional efforts are needed to reach older workers – the demographic group most likely to believe rumors within our sample.

Finally, participants in our study reported high confidence in the government. This finding was unexpected, because: (i) previous studies had linked the spread of rumors to low trust in government [29], (ii) the pandemic had revealed health disparities between participants and the general population [17], and (iii) a non-trivial group of participants (40%) expressed fears about their health or about job security. Nonetheless, interviews with migrant workers suggest that they may have compared their situation to that of other countries [30]. As Singapore’s COVID-19 response is perceived as ‘high performing’ on the global scale [21, 31], the government could have gained participants’ confidence in this manner. We thus urge further research to better understand the views of migrant workers across different countries.

In reporting these findings, we note several limitations. First, we only captured migrant workers’ responses at one time-point. As the COVID-19 situation is fluid, more studies are needed to understand how changing circumstances (e.g., the emergence of new variants, vaccination programs) influence responses. Second, our survey used self-reported measures. These measures may be subject to recollection biases or social pressures (e.g., fear of repercussions within a host country), and follow-up research can consider alternate data sources (e.g., mining social media posts).

## 5. Conclusions

To conclude, our study provides a rare window into migrant workers’ views amidst large COVID-19 outbreaks. Despite being confident in the government’s response, participants in our study showed a high reliance on COVID-19 rumors. These findings provide a guideline for public health policies addressing migrant worker communities.

## Data Availability

As per the IRB protocol, data will be available upon request.

## Author contributions

JCJL, PB and KD conceived of the study. EYQT, DA, YES and JCJL carried out data collection and data analyses, accessed and verified the underlying data, and wrote the first draft of the manuscript. All authors contributed to editing and commenting on the final version.

## Acknowledgements

We are grateful to the migrant worker community who hosted us; the surveyors and translators who carried out data collection; hospital, government, and facility staff who granted us research access; and the HealthServe team for feedback during the design phase.

